# Kidney stone prevalence based on self-report and Electronic Health Records: insight into the prevalence of active medical care for kidney stones

**DOI:** 10.1101/2022.05.17.22275212

**Authors:** Connor M. Forbes, Naren Nimmagadda, Nicholas L. Kavoussi, Yaomin Xu, Cosmin A. Bejan, Nicole L. Miller, Ryan S. Hsi

## Abstract

**Introduction:** Kidney stone prevalence estimates vary depending on sampling methodology. We compared rates of patient-reported kidney stone disease to Electronic Health Records (EHR) kidney stone diagnosis using a common dataset to evaluate for socio-demographic differences in these populations, including between those with and without active care for kidney stones.

**Methods:** From the All of Us research database, we identified 21,687 adult participants with both patient-reported medical history and EHR data. We extracted patient-reported kidney stone history and medical encounters for kidney stones from EHR. We compared differences in age, sex, race, education, employment status and healthcare access between patients with self-reported kidney stone history without EHR data to those with EHR-based diagnoses.

**Results:** In this population, the self-reported prevalence of kidney stones was 8.6% overall (n=1877), including 4.6% (n=1004) who had self-reported diagnoses but no EHR data. Among those with self-reported kidney stone diagnoses only, the median age was 66, 43% were male, and 92% were Non-Hispanic Whites, compared 120,623 (53.9%) in the entire All of Us cohort. The EHR-based prevalence of kidney stones was 5.7% (n=1231), median age 67, of whom 45% were male and 92% were Non-Hispanic White. No differences were observed in age, sex, education, employment status, rural/urban status, or ability to afford healthcare between groups with EHR diagnosis or self-reported diagnosis only. Of patients who had a self-reported history of kidney stones, 24% reported actively seeing a provider for kidney stones.

**Conclusions:** Kidney stone prevalence by self-report is higher than EHR-based prevalence in this national dataset. Using either method alone to estimate kidney stone prevalence may exclude some patients with the condition, although the demographic profile of both groups is similar. Approximately one in four patients report actively seeing a provider for stone disease.

## Introduction

Kidney stone prevalence estimates have been determined either by self-report or using electronic databases,^1-3^ however, these methodologies yield different estimates of disease prevalence. Accurately determining disease prevalence has wide-ranging implications, including directing policy priorities, developing of clinical guidelines, providing the clinical context for diagnostic decision making,^4^ and estimating the cost burden of care.^5^

Reconciling estimates derived from self-reported and electronic databases for kidney stone prevalence has been difficult due to differences in study design, geography/climate over time,^6,7^ and changing demographics of the population^8^. Additionally, while it is known that not all patients with kidney stones seek medical care, the accuracy of self-reported prevalence is challenging to ascertain. It is unknown whether certain patients are systematically excluded from one sampling method, such as patients who may have limited access to care or have lower socioeconomic status.

Within this context, we investigated a research database in the United States focused on the recruitment of underrepresented individuals in medical research, which includes both self-reported and Electronic Health Records (EHR) diagnoses of kidney stones. We compared characteristics of these patient populations based on two prevalence definitions to describe patients who were omitted from either method, and characterized those who reported actively receiving medical care for kidney stone disease.

## Methods

### Study population

We examined the “All of Us” research database, which is an National Institute of Health funded database drawn from participants across the United States, with a recruitment emphasis from populations who are traditionally underrepresented in biomedical research.^9^ To this end, the program funds 22 community partners to encourage recruitment.^10^ The initial available cohort includes more than 220,000 patients, with recruitment ongoing towards a goal of 1 million participants.^9^ This study was approved by the All of Us Institutional Review Board and complies with reporting requirements, including avoiding reporting data with <20 patients to preserve anonymity. Here, the study cohort examined included participants recruited from 2016 to 2019 (n=223,921), of whom 61% were female (n=136,223), 53.9% White (n=120,623), 20.8% Black/African American (n=46,655), and 3.3% Asian (n=7,485). Then, we restricted the population to those with both self-reported medical history and EHR data available for analysis, yielding 21,687 participants.

### Self Reported Kidney Stone History and Active Care

Self-reported personal medical history surveys were administered from 2016 to 2019 at the time of recruitment, with all recruited participants given the opportunity to participate. Of recruited patients, 39,200 had survey data available defined as providing at least one answer. In addition to comprehensive medical history data, patients were asked: “Has a doctor or health care provider ever told you that you have any of the following?” and were provided a list of options. Patients who selected “Yes” for “Kidney stones” were included in the self-reported diagnosis of kidney stones population. In the same survey, patients were asked “Are you still seeing a doctor or health care provider for…” and were categorized as having self-reported active clinical care if they selected “Yes” for “Kidney stones.” Self-report of kidney stone episodes has been found in a prior study to have a validity of 97%.^11^

### Kidney Stone EHR Diagnosis

SNOMED and ICD condition and procedure codes were used to identify kidney stone diagnoses and procedures (see Appendix I). These included condition codes related to kidney and ureteral stones, and procedure codes related to kidney shockwave lithotripsy, ureteroscopy with stone treatment, and percutaneous nephrolithotomy. The allowable time period for these codes was throughout Electronic Health Record (EHR) data availability, which ranged from a first code start date of 1980 to 2017. Patients were categorized as having a kidney stone if they had 1 or more condition or procedure codes. A prior study examining the validity of ICD codes for kidney stone disease had a positive predictive value of 96%.^3^

### Covariates

Covariates included age, sex, race, education level, employment status, and total household income. Age was calculated as current age at time of analysis. Sex was based on survey response to the question: “What was your assigned sex at birth?” Education level, employment status, and total household income were based on survey responses. Barriers to healthcare including ability to afford specialist care and rural location with long distance to healthcare provider were assessed through survey questions.

### Statistical analysis

We categorized distinct kidney stone patient cohorts based on the presence of self-reported and EHR-based definitions of kidney stone disease (see Figure 1). First, to evaluate whether patients with kidney stones identified by EHR were distinct from patients with self reported kidney stones, we compared patients who were diagnosed as having a kidney stone by EHR data (Figure 1, Cohort A) to those who were only diagnosed by self-report and without associated EHR data (Figure 1, Cohort B). Then, to evaluate whether patients receiving active medical care were different from those without, we examined patients who had survey data available with self-reported kidney stones (Figure 1, Cohort C), and compared socio-demographic differences among patients reporting active medical care for kidney stones versus those without. For both analyses we compared age, sex, race, education, employment status and healthcare access using the Mann-Whitney U test for continuous variables and Pearson’s Chi-squared test for categorical variables. Per All of Us policy, no published data can contain fewer than 20 participants in a single cell. In some cases, such as race, this necessitated combining categories. We designated alpha for all statistical tests as 0.05 for statistical significance. Python software (v3.7.12, 2021) was used for the analysis.

**Figure 1.**
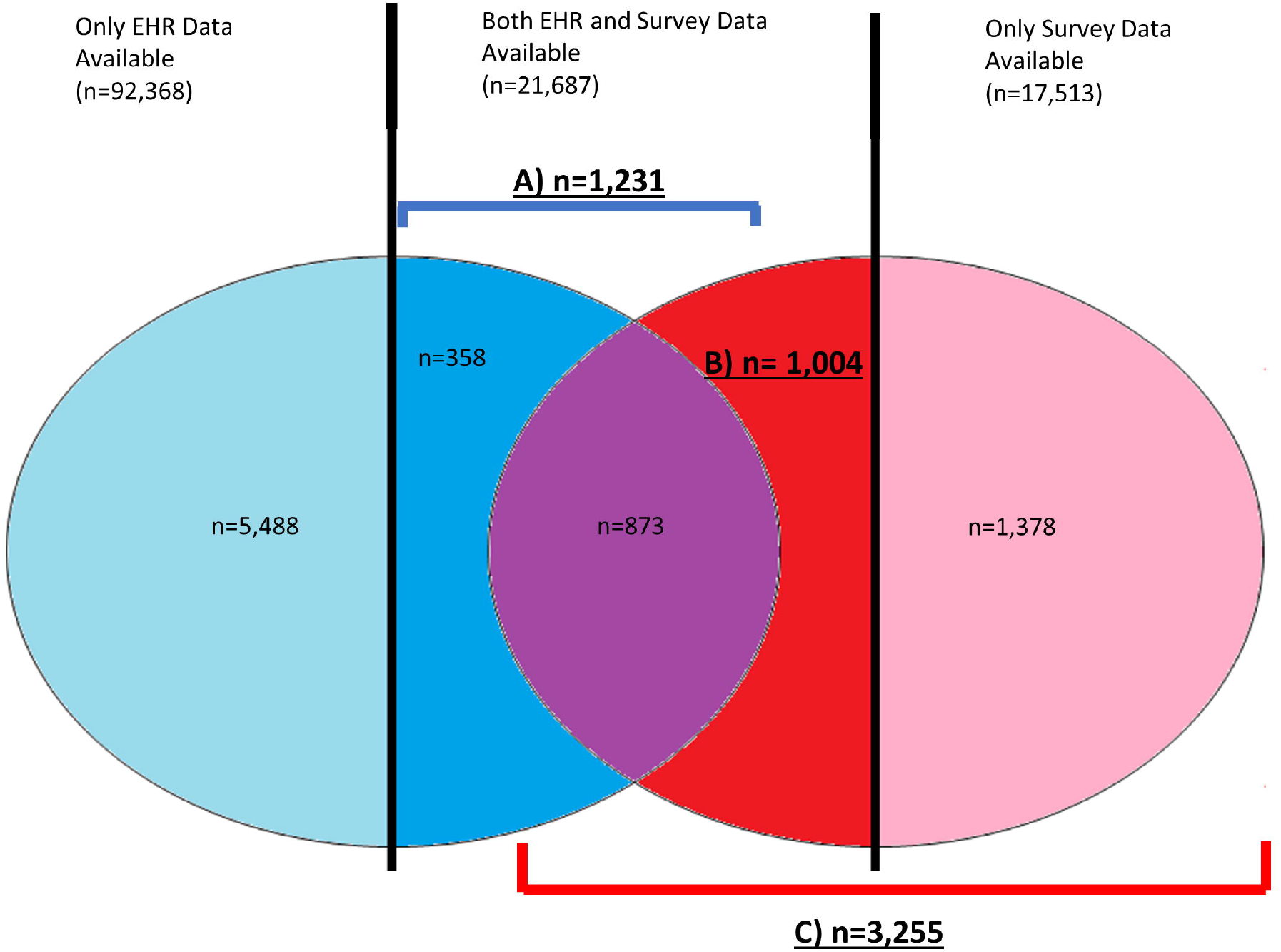
Electronic Health Records (EHR) based prevalence (blue circle) and self reported prevalence (red circle) of kidney stones. Groups chosen for analysis are in labelled and bold. A) (dark blue and purple) EHR based prevalence of kidney stone disease, restricted to patients with both EHR and survey data available (n = 1231). B) (red only) self-reported prevalence of kidney stones only, restricted to patients with both EHR and survey data available (n=1004). C) (purple, red and pink) total self reported prevalence of kidney stones. A/B are sources for Table 1; C is source for Table 2.

## Results

Of 223,921 participants in All of Us, there were 114,055 patients with EHR data available. Of these, 6,716 (5.9%) patients had an EHR-based diagnosis of kidney stones. Of 39,200 patients who completed a personal medical history survey, there were 3,255 (8.3%) with a self-reported history of kidney stones. When restricted to patients who had both survey and EHR data available (n=21,687), there were 1231 patients (5.7%, Cohort A, Figure 1) with an EHR diagnosis of kidney stones and 1877 (8.6%) had a self-reported diagnosis of kidney stones.

There were 1004 patients with a self-reported diagnosis only, without an associated EHR diagnosis (4.6% of those with both EHR and survey data available, Cohort “B”). This represents 45% of the total number of patients with all data available with a diagnosis of kidney stones. The median age of the groups was 67 for the EHR-based diagnosis group and 66 for the self-reported diagnosis only group (p=0.07). No differences were observed in sex, race, education, employment status, rural location, or ability to afford specialist care or follow up care (Table 1). Despite being drawn from patients who had EHR data available, Cohort B did not have non-kidney stone conditions/procedure codes after excluding those with kidney stone conditions/procedure codes.

**Table 1.**
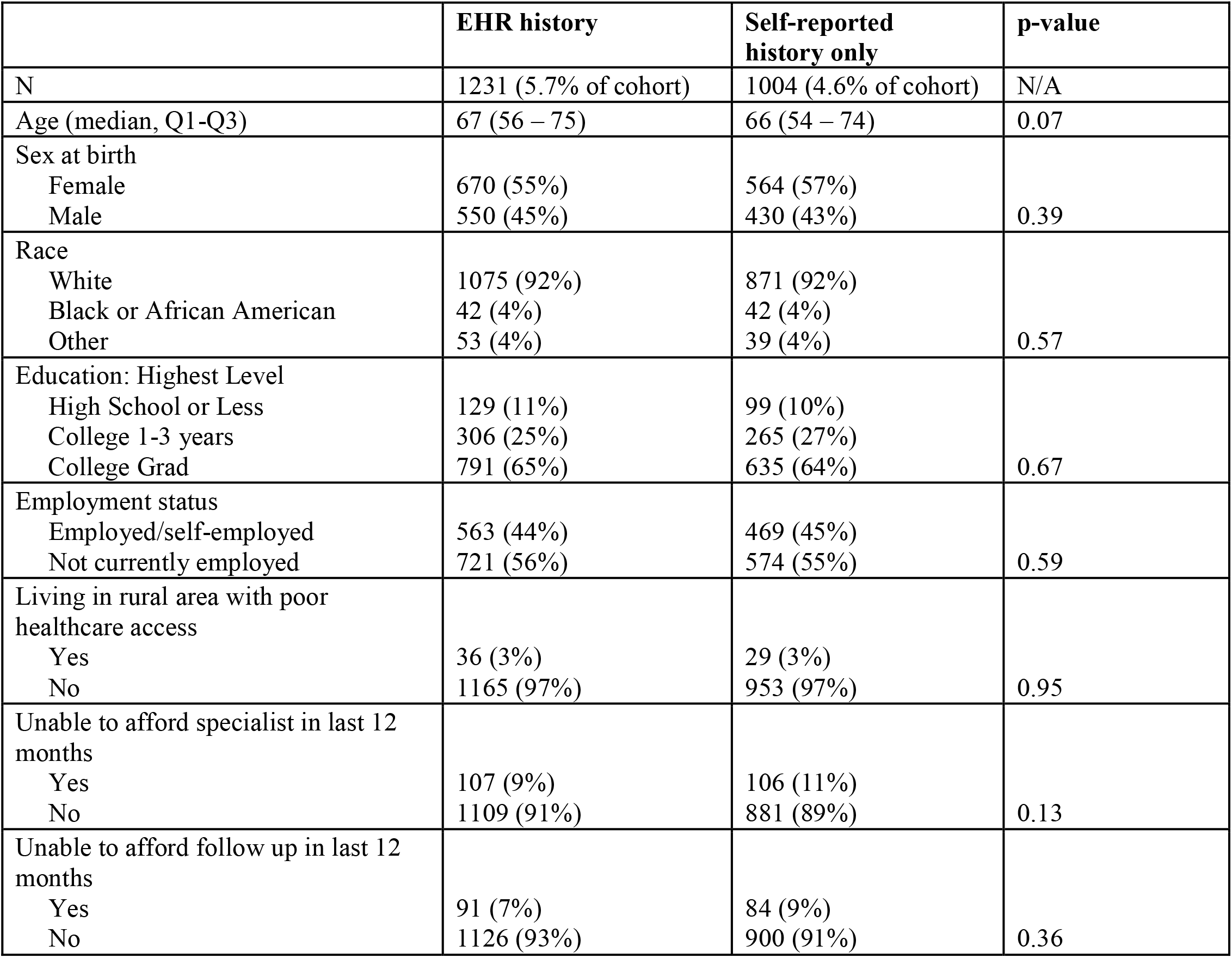
Demographics of kidney stone patients diagnosed by Electronic Health Records (EHR) or by patient-report alone. Left column is cohort “A” and right column is cohort “B” from Figure 1. Q1 = 1^st^ Quartile; Q3 = 3^rd^ Quartile.

We then assessed the cohort of 3,255 patients who had a self-reported history of kidney stones and compared those who did or did not report actively seeing a provider for kidney stone care (Table 2). Of these, 782 patients (24%) reported actively seeing a provider for kidney stones. Patients who were actively seeing a physician were significantly older (67 vs 65, p=0.04). Significantly more patients who were actively seeing a provider were male compared to those who were not actively seeing a provider (n=395, 51% vs n=1130 male, 46%, p=0.02). Fewer patients who were actively seeing a provider had advanced college degrees compared to those who were not actively seeing a provider (n=506, 65% vs n=1686, 68%, p=0.02). Fewer patients who were actively seeing a provider were employed (n=335, 41%, vs n=1290, 50%, p<0.001), and more patients who were actively seeing a provider had an associated kidney stone-related EHR diagnosis recorded (n=352, 45% vs n=514, 21%, p<0.001). No differences were observed in race, rural location, reported income, or reported ability to afford specialist or follow up care (Table 2).

**Table 2.** Patients with kidney stones (by self report) who report actively seeing a physician compared to those who do not. These patients are drawn from cohort “C” in Figure 1. EHR = Electronic Health Record Q1 = 1^st^ Quartile; Q3 = 3^rd^ Quartile.

|  | Actively seeing physician<br>for kidney stones | Not actively seeing<br>physician for kidney<br>stones | p-value |
| --- | --- | --- | --- |
| N | 782 (24%) | 2473 (76%) | N/A |
| Age (median, Q1-Q3) | 67 (55 – 74) | 65 (53 – 73) | 0.04* |
| Sex at birth |  |  |  |
| Female | 380 (49%) | 1326 (54%) |  |
| Male | 395 (51%) | 1130 (46%) | 0.02* |
| Race |  |  |  |
| White | 699 (94%) | 2216 (93%) |  |
| Non-white | 47 (6%) | 167 (7%) | 0.45 |
| Education: Highest Level |  |  |  |
| High School or Less | 79 (10%) | 175 (7%) |  |
| College 1-3 years | 193 (25%) | 604 (25%) |  |
| College Grad/Advanced Degree | 506 (65%) | 1686 (68%) | 0.02* |
| Employment status |  |  |  |
| Employed/self-employed | 335 (41%) | 1290 (50%) |  |
| Not currently employed | 486 (59%) | 1300 (50%) | <0.001** |
| Living in rural area with poor<br>healthcare access |  |  |  |
| Yes | 32 (4%) | 82 (3%) |  |
| No | 728 (96%) | 2339 (97%) | 0.29 |
| Unable to afford specialist in last 12<br>months |  |  |  |
| Yes | 82 (11%) | 252 (10%) |  |
| No | 689 (89%) | 2182 (90%) | 0.82 |
| Unable to afford follow up in last 12<br>months |  |  |  |
| Yes | 61 (8%) | 196 (8%) |  |
| No | 708 (92%) | 2238 (92%) | 0.91 |
| Total household income, n (%) |  |  |  |
| <\$50,000 | 210 (30%) | 634 (28%) | |
| \$50,000-\$100,000 | 219 (31%) | 736 (32%) | |
| >\$100,000 | 275 (39%) | 916 (40%) | 0.56 |
| Patients with a prior kidney stone-related EHR diagnosis, n (%) |  |  |  |
| Yes | 352 (45%) | 514 (21%) |  |
| No | 430 (55%) | 1959 (79%) | <0.001* |

## Discussion

In this study, we evaluated two methods of kidney stone prevalence estimation in the same population. We found different prevalence estimates of kidney stones using patient self report and EHR data (8.6% versus 5.7%, respectively). We found that almost half of the patients with kidneys stones were omitted if using an EHR diagnosis only. Only 1 in 4 of the self-diagnosed cohort reported actively seeing a provider for kidney stones. These prevalence rates compare similarly to previous literature in different populations: a prior study examining self-reported diagnoses using the U.S.-based National Health and Nutritional Examination Survey (NHANES) (11%),^8^ and a database study reporting a prevalence of 3.5% in a different population.^2^

Recent evaluations of kidney stone occurrence have found an increase in non-white patients with stones. The odds ratios for non-white patients for kidney stones ranged from 0.37 – 0.60 based on data from 2007-2010,^1^ which increased to 0.51 – 1.06 in 2015 – 2018.^8^ The percentage of non-white patients diagnosed with kidney stones in the NHANES national survey was 27.1%.^8^ Notably, our cohort with diagnoses of kidney stones was >90% white, whereas the overall cohort was 54% white. Our study findings compared to that of the NHANES national survey likely reflect distinct sampling methods and further illustrate the need to investigate whether these time trends showing increasing kidney stone prevalence among minorities continue.

Lack of adherence with treatment recommendations and loss to follow up is a known issue for kidney stone disease management.^12,13^ Since active follow up and preventive interventions reduce recurrence,^14,15^ the high number of patients who are not receiving active care – 3 of every 4 patients in our study – represent an area for investigation to determine if this is appropriate care or a missed opportunity for disease trajectory modification. Demographic differences in the group not receiving active care included female preponderance, younger age, advanced college degree attainment, and active employment. It is possible that these patients had more difficulty scheduling medical appointments due to work-related issues^16^ or had barriers to medical care based on social dynamics. Addressing any possible barriers to care in kidney stone patients may increase engagement and requires additional study.

Do all patients with kidney stone disease need active medical management? Previous data suggests that lower socioeconomic status is associated with an increased stone burden at time of referral or intervention.^17,18^ We had therefore hypothesized that patients who self-reported stones but did not have associated EHR diagnosis might be deferring care for reasons of cost or limited access. However, we could not detect differences in demographic factors, including race, employment status, rural location, or ability to afford medical care. It is also possible that these individuals may have EHR diagnoses not captured in our dataset, or self-diagnosed the condition outside of the health care system. This suggests that the prevalence of kidney stones may be systematically underestimated using condition/procedure codes alone. A recent study of the All of Us research program overall found that strength of concordance between EHR-based diagnosis and survey data for diseases vary.^19^ The authors suggest that survey data overall provides a useful avenue to augment EHR data through identifying missing records or diagnoses, and addressing biases in EHR data collection.

The limitations of this study include the potential for missing data, inaccurate coding and recall bias. In this dataset, more patients who reported seeing a provider for stone care had an associated EHR diagnosis (45% versus 21%, p<0.001). However, not all of these patients had EHR data available for this cohort. It is possible that the patients with self-reported stones had a remote history of kidney stone disease prior to the available EHR data, or that there were gaps in their available EHR data at the time of their kidney stone treatment. For example, our cohort of self-diagnosis of kidney stones without associated kidney stone EHR diagnosis did not have other condition/procedure codes present, although their EHR data contained laboratory and imaging values. This suggests the possibility that their data were less complete. However, a prior publication examining the concordance of self-reported and EHR data in All of Us showed that traditionally underreported EHR diagnoses such as myopia are low, while traditionally highly reported diagnoses such as cancer are high, suggesting that the data are appropriately collected.^19^ Furthermore, issues with missing data could represent a systematic bias in EHR based kidney stone prevalence assessment globally. Finally, income and education were used as surrogates for financial ability, but net worth, which is likely a better indicator of socioeconomic status in this older population, could not be assessed.

## Conclusions

In this national database, prevalence estimates of kidney stone disease differed when using self-report or EHR data. Either method alone omitted a large number of individuals. The limitations of these sampling methodologies should be considered when extrapolating for policy and guideline decisions. The majority of patients with a history of kidney stones do not receiving active care for prevention, which may be a targetable area for improvement. Those less likely to receive active medical care are slightly younger, more often female, have an advanced college degree, and have active employment.

## Data Availability

All data produced in the present study are available upon reasonable request to the authors

## Acknowledgements

This project utilized the All of Us research database. The All of Us Research Program is supported by the National Institutes of Health, Office of the Director: Regional Medical Centers: 1 OT2 OD026549; 1 OT2 OD026554; 1 OT2 OD026557; 1 OT2 OD026556; 1 OT2 OD026550; 1 OT2 OD 026552; 1 OT2 OD026553; 1 OT2 OD026548; 1 OT2 OD026551; 1 OT2 OD026555; IAA #: AOD 16037; Federally Qualified Health Centers: HHSN 263201600085U; Data and Research Center: 5 U2C OD023196; Biobank: 1 U24 OD023121; The Participant Center: U24 OD023176; Participant Technology Systems Center: 1 U24 OD023163; Communications and Engagement: 3 OT2 OD023205; 3 OT2 OD023206; and Community Partners: 1 OT2 OD025277; 3 OT2 OD025315; 1 OT2 OD025337; 1 OT2 OD025276. In addition, the All of Us Research Program would not be possible without the partnership of its participants. RH, YX, and CB are supported by R21 DK127075.

## Author Statements

All authors certify that they had substantial contributions to the manuscript, including conception/data acquisition/analysis; drafting/critical revision of the work; final approval of the work; agreement of accountability of the work. Specific work includes the following roles. CMF: Formal Analysis, Conceptualization, Writing – Original Draft; NN: Conceptualization, Project Administration, Writing – Review and Editing; NK: Visualization, Writing – Review and Editing; YX: Formal Analysis, Writing – Review and Editing; CAB: Formal Analysis, Methodology, Writing – Review and Editing; NLM: Conceptualization, Supervision, Resources. Writing – Review and Editing; RSH: Conceptualization, Project Administration, Resources, Supervision, Writing – Original Draft.

Author conflict of interest: the authors have no relevant conflicts of interest to disclose.

