## Appendix I for "Kidney stone prevalence based on self-report and Electronic Health Records: insight into the prevalence of active medical care for kidney stones"

**Appendix I. Condition and procedure codes used for diagnosis of kidney/ureteral stones.**

**Condition codes:**

"Kidney Stone":

SNOMED 95570007

ICD10CM-N20.0

ICD10-N20.0

ICD9CM-592.0

SNOMED 95570007

SNOMED 56491003

OXMIS-592 N

"Ureteric stone":

ICD9CM-592.1

ICD10CM-N20.1

SNOMED-31054009

ICD10-N20.1

SNOMED-95573009

"Calculus of the kidney and ureter"

ICD10CM-N20

ICD9CM-592

ICD10CM-N20.2

SNOMED-266556005

ICD10-N20.2

“Hydronephrosis co-occurrent and due to calculus of the kidney and ureter":

ICD10CM-N13.2

SNOMED-736640009

ICD10-N13.2

"calyceal renal calculus":

SNOMED 236708007

**Procedure codes:**

“Lithotripsy, extracorporeal shock wave on the kidney”

CPT 50590

“Cystourethroscopy, with ureteroscopy and/or pyeloscopy; with lithotripsy including insertion of indwelling ureteral stent”

CPT 52356

"Percutaneous extraction of kidney stone with fragmentation procedure":

SNOMED-42041003

ICD9Proc-55.04

"Percutaneous nephrostolithotomy or pyelostolithotomy, with or without dilation, endoscopy, lithotripsy, stenting, or basket extraction; up to 2 cm":

CPT4-50080

"Percutaneous nephrostolithotomy or pyelostolithotomy, with or without dilation, endoscopy, lithotripsy, stenting, or basket extraction; over 2 cm":

CPT4-50081

"Cystourethroscopy, with ureteroscopy and/or pyeloscopy; with lithotripsy (ureteral catheterization is included)":

CPT4-52353

"Cystourethroscopy, with ureteroscopy and/or pyeloscopy; with removal or manipulation of calculus (ureteral catheterization is included)"
CPT4-52352

"Cystourethroscopy, with ureteroscopy and/or pyeloscopy; with endoscopic laser treatment of ureteral calculi (includes ureteral catheterization)"

HCPCS-S2070

"Renal endoscopy through established nephrostomy or pyelostomy, with or without irrigation, instillation, or ureteropyelography, exclusive of radiologic service; with removal of foreign body or calculus"
CPT4-50561

"Extracorporeal shockwave lithotripsy [ESWL] of the kidney, ureter and/or bladder"

ICD9Proc-98.51

"Extracorporeal shockwave lithotripsy of the kidney"

SNOMED-24376003

ICD9Proc-59.95

"Pyelolithotomy"

SNOMED-36732002

"Nephrolithotomy; removal of large staghorn calculus filling renal pelvis and calyces (including anatrophic pyelolithotomy)"

CPT4-50075

"Pyelotomy; with removal of calculus (pyelolithotomy, pelviolithotomy, including coagulum pyelolithotomy)"

CPT4-50130
